# Incremental Predictive Value and Representation of Transcriptomic Features for Immunotherapy Response in Advanced Urothelial Carcinoma

**DOI:** 10.64898/2026.09.14.26363060

**Authors:** Grace Y. Han, Hansen Tai, Fusheng Wang

## Abstract

Transcriptomic profiling may improve immunotherapy response prediction, but its incremental value beyond established clinical and tumor biomarkers is unclear. In 298 patients with advanced urothelial carcinoma from IMvigor210, an atezolizumab (PD-L1 inhibitor) trial, we compared knowledge-guided tumor microenvironment gene-set features with data-driven transcriptome-wide gene selection. Knowledge-guided features improved AUPRC by 0.058 beyond clinical variables alone, with smaller gains after including standard biomarkers (TMB, PD-L1; +0.011) and research-enriched biomarkers (neoantigen burden, immune phenotype; +0.016). Data-driven gene selection was unstable across folds (mean Jaccard similarity, 0.227) and, under the clinical-plus-biomarker baseline, yielded an AUPRC 0.150 lower than the knowledge-guided representation. Despite this instability, IL-6/JAK/STAT3 signaling was the most recurrent Hallmark pathway, suggesting pathway-level biological convergence. External evaluation showed a similar pattern: knowledge-guided features improved AUPRC by 0.128 over clinical variables vs. 0.019 over clinical-plus-biomarker baseline. These findings support biologically informed transcriptomic representation when molecular profiling adds value beyond established biomarkers.

## Introduction

Immune checkpoint inhibitors (ICIs) targeting programmed cell death protein 1 (PD-1) or programmed death ligand 1 (PD-L1) have transformed the treatment of advanced urothelial carcinoma by restoring antitumor T-cell activity. However, durable responses occur in only a subset of patients. Across studies of single-agent PD-1/PD-L1 blockade in locally advanced or metastatic urothelial carcinoma (mUC), objective response rates have generally remained between approximately 20% and 30%, depending on treatment setting and patient population.^1, 2^ Because ICIs can also cause clinically significant immune-related toxicities and impose treatment burden, this heterogeneity creates a critical unmet need for precision strategies that more reliably identify patients likely to respond.

The heterogeneous response to ICIs has motivated extensive efforts to identify predictive biomarkers. PD-L1 expression and tumor mutational burden (TMB) are among the most established and clinically measurable biomarkers associated with ICI response, but neither provides sufficient discrimination for reliable patient stratification. PD-L1 reflects engagement of an immunosuppressive checkpoint pathway but is limited by spatial heterogeneity, assay dependence, and dynamic expression, whereas TMB approximates tumor immunogenicity through mutational load but does not capture whether an effective antitumor immune response is present^3^. Tumor neoantigen burden provides a related measure of potentially immunogenic mutation-derived peptides, but antigenicity alone is insufficient to generate effective tumor control. Response also depends on the tumor microenvironment (TME), where a complex and dynamic interplay of tumor and host factors (e.g., immune infiltration, stromal interactions, vascualr context) can promote immune activation, suppression, or exclusion. TME phenotypes characterized as inflamed, excluded, or desert therefore capture dimensions of tumor-immune biology not fully represented by tumor-intrinsic biomarkers^4^.

Transcriptomic profiling provides a molecular readout of these coordinated tumor-immune processes. Interferon-γ-related and T-cell-inflamed gene-expression programs have been associated with response to PD-1/PD-L1 blockade, whereas stromal programs associated with immune exclusion have been linked to resistance^4, 5^. Tumor mutational and T-cell-inflamed gene-expression signals capture distinct but related dimensions of ICI response, reflecting tumor immunogenicity and an active antitumor immune state, respectively^6^. These findings establish the biological relevance of transcriptomic features, but biological association does not establish how much predictive information they contribute beyond clinical characteristics and tumor biomarkers already available to a prediction model.

Recent computational studies have demonstrated the predictive potential of transcriptomic features in mUC. Piyawajanusorn et al. developed gene-expression models for atezolizumab response and compared them with models incorporating clinical and tumor biomarker features^7^. Langfelder et al. derived a 49-gene signature that showed external predictive performance and was evaluated against PD-L1, established immune-expression signatures, and TMB-based models^8^. Together, these studies established that transcriptomic signatures can support response prediction, but their primary focus was signature development and comparative predictive performance. What remains unclear is how much transcriptomics adds beyond increasingly comprehensive clinical and tumor biomarker baselines. A related question is how transcriptomic information should be represented. In clinical transcriptomic datasets, high dimensionality and limited sample size can make gene-level feature selection sensitive to sampling variability^9^. Whether biologically guided aggregation provides a more stable and predictive representation than transcriptome-wide gene selection under the same modeling framework also remains unclear. As sequencing becomes increasingly accessible, the translational informatics challenge is therefore not simply whether transcriptomic data can predict response, but when they add information beyond existing biomarkers and how to best represent the data to support precision immuno-oncology.

In this study, we evaluated pretreatment transcriptomic features for predicting response to PD-L1 blockade in mUC. Our study makes three contributions. First, it quantified the incremental predictive value of knowledge-guided TME gene-set scores across progressively richer clinical and tumor biomarker baselines. Second, it compared this biologically informed representation with data-driven transcriptome-wide gene selection. Third, it evaluated whether the relative predictive pattern was reproducible in an independent cohort of ICI-treated patients. By evaluating both transcriptomic representations under a common predictive framework, our study provides a general framework for evaluating the addition of high-dimensional molecular data to multimodal clinical prediction models.

## Methods

### Dataset and Study Cohort

We used the IMvigor210 dataset, a Phase II, single-arm clinical trial evaluating atezolizumab (1,200 mg intravenously every 21 days), a PD-L1 inhibitor, in patients with locally advanced or metastatic urothelial carcinoma^1^. The dataset is publicly available through the `IMvigor210CoreBiologies` R package^4^ and contains matched clinical, biomarker, and transcriptomic data for 348 patients. Tumor response was assessed according to Response Evaluation Criteria in Solid Tumors (RECIST) version 1.1. Patients without an evaluable response were excluded from response modeling. Complete response (CR) and partial response (PR) were classified as responders, whereas stable disease (SD) and progressive disease (PD) were classified as non-responders.

Predictors were organized into clinical, standard biomarker, research-enriched biomarker, and transcriptomic feature blocks. The clinical block (*C*) included sex, Eastern Cooperative Oncology Group (ECOG) performance status, tobacco use history, metastatic disease site, prior platinum therapy, and prior intravesical bacillus Calmette-Guérin (BCG) therapy. The standard biomarker block (*B*) included tumor mutational burden (TMB) and immune-cell (IC) and tumor-cell (TC) PD-L1 expression measured by SP142 immunohistochemistry (IHC). The research-enriched biomarker block (*R*) included neoantigen burden and immune phenotype, categorized as inflamed, excluded, or desert. Pretreatment RNA-sequencing profiles spanning 31,286 genes were used to derive transcriptomic features.

### Transcriptomic Feature Engineering

Transcriptomic features were engineered using two complementary dimensionality-reduction strategies. The knowledge-guided approach summarized gene expression using prespecified TME gene sets, and the data-driven approach learned a sparse set of predictive genes using elastic-net selection. Raw RNA-sequencing counts were converted to transcripts per million (TPM) using gene lengths and transformed as log_2_(TPM + 1) before feature engineering.

#### Knowledge-Guided TME Gene Sets

Eight gene sets were selected from prior literature to capture key TME processes spanning immune activation, antigen presentation, immune regulation, myeloid infiltration, and stromal and vascular biology. These included CD8 T-effector (8 genes), immune checkpoint (7 genes), antigen-processing machinery (APM; 6 genes), natural killer (NK) cells (9 genes), cytotoxic lymphocytes (7 genes), monocytic lineage (7 genes), fibroblasts (8 genes), and endothelial cells (33 genes)^4, 10^. To provide a consistent representation, each gene-set score was calculated as the mean log_2_ (TPM + 1) expression of its constituent genes. This transformation reduced 31,286 gene-level measurements to eight biologically interpretable features, which collectively comprised the knowledge-guided transcriptomic block (*K*). Individual gene-set scores were compared between responders and non-responders using two-sided Mann–Whitney U tests. For each gene set, median scores in each response group and the difference in medians (responders minus non-responders) were reported. Benjamini–Hochberg false discovery rate (BH-FDR) correction was applied across the eight comparisons, with *q* < 0.05 considered statistically significant^11^.

#### Data-Driven Feature Selection

A separate data-driven approach was used to derive a high-dimensional transcriptomic representation for comparison with the prespecified knowledge-guided representation. Feature selection was performed independently within each outer training set of the repeated cross-validation (CV) framework (see *Model Development* below). Genes with variance ≤ 0.05 in log_2_ (TPM +1) expression were first excluded. Elastic-net penalized logistic regression was then applied to the remaining genes, with the *l*_1_ ratio selected from {0.5, 0.7, 0.9, 1.0} and inverse regularization strength from {0.01, 0.1} using stratified three-fold CV and average precision as the optimization metric^12^. Genes with nonzero coefficients in the selected elastic-net model were retained. Variance filtering, elastic-net tuning, and gene selection were performed using only the corresponding outer training set. Because this supervised, fold-specific approach could select different genes across training samples, stability was assessed using selection frequency and pairwise Jaccard similarity across the 50 outer folds. Jaccard similarity was defined as the proportion of shared genes relative to the union of two selected sets, with higher values indicating greater overlap.

To determine whether fold-specific gene selections converged on common biological pathways, overrepresentation analysis was performed using the 50 Molecular Signatures Database (MSigDB) Hallmark gene sets^13^. Within each outer training set, selected genes were evaluated using a one-sided hypergeometric test, with genes that passed the variance filter in that training set serving as the background universe. BH-FDR correction was applied across the 50 pathways within each fold, with *q* < 0.05 defining significant enrichment. The five pathways with the smallest enrichment *p*-values in each fold were also recorded irrespective of statistical significance, and their recurrence across the 50 outer fits was summarized.

### Model Development

Two complementary modeling analyses were performed. First, the biomarker-richness analysis evaluated whether knowledge-guided transcriptomic features provided incremental predictive information as progressively richer non-transcriptomic information became available. The following comparisons were examined: clinical variables alone vs. clinical plus knowledge-guided transcriptomic features (*C* vs. *C* + *K*); clinical plus standard biomarkers vs. clinical plus standard biomarkers and knowledge-guided transcriptomic features (*C* + *B* vs. *C* + *B* + *K*); and clinical, standard biomarker, and research-enriched biomarker features vs. the corresponding model with knowledge-guided transcriptomic features (*C* + *B* + *R* vs. *C* + *B* + *R* + *K*). The *C* + *B* vs. *C* + *B* + *K* comparison was designated as the primary comparison because it most closely reflects information routinely available in clinical practice, whereas Block *R* was considered research-enriched because neoantigen burden and immune phenotype are not routinely available as standard clinical tests. Second, the transcriptomic-representation analysis compared the knowledge-guided and data-driven transcriptomic representations under the clinical plus standard biomarker (*C* + *B*) baseline. The following models were evaluated: *C* + *B, C* + *B* + *K*, and *C* + *B* + elastic-net-selected genes, with elastic-net-selected genes determined independently within each outer training set as described above.

Ridge (*L*_2_-penalized) logistic regression was used for all final predictive models^14^. The *L*_2_ penalty was well suited to the expected correlation among the prespecified clinical, biomarker, and transcriptomic features. Model coefficients were estimated by minimizing the penalized negative log-likelihood, 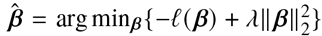, where *ℓ*(***β***) denotes the logistic log-likelihood and *λ* controls the strength of *L*_2_ regularization. By shrinking coefficient magnitudes toward zero while retaining all predictors, ridge regularization reduces variance in the presence of correlated features and allows them to contribute jointly without imposing additional feature selection. Given the modest cohort size and limited number of responders, a regularized linear model was also favored over more flexible algorithms to reduce model variance and the risk of overfitting. Logistic regression additionally provides predicted response probabilities, enabling evaluation of both discrimination and probabilistic performance. The same model class was used throughout so that performance differences reflected differences in feature information rather than differences between learning algorithms. No class weighting or resampling was applied because these approaches can impair probability calibration without improving discrimination in logistic prediction models^15^.

Final ridge models were trained and tuned using stratified nested five-fold CV repeated 10 times. In each outer fold, approximately 80% of patients were used for training and 20% for held-out evaluation. Within the training set, preprocessing and model fitting were implemented in a single pipeline to prevent information leakage. Continuous predictors were median-imputed with missingness indicators and standardized; categorical predictors were assigned a missing category and one-hot encoded. Five-fold inner CV selected the inverse regularization strength from 0.001, 0.01, 0.1, 1, 10, 100, 1000 using average precision. The model was then refit on the full outer training set and applied to the held-out patients. All preprocessing and tuning parameters were estimated from training data only.

### Model Evaluation

Held-out predictions from the five outer folds were pooled within each repeat to obtain one out-of-fold prediction per patient per repeat. Area under the precision–recall curve (AUPRC) was the primary performance metric because responders comprised 22.8% of the cohort^16^. Area under the receiver operating characteristic curve (AUROC) and Brier score were also evaluated. For the biomarker-richness analysis, incremental predictive value was defined as the paired difference between each model including Block *K* and its corresponding baseline. For example, ΔAUPRC = AUPRC _+*K*_ − AUPRC_baseline_. The same convention was used for AUROC and Brier score; positive ΔAUPRC and ΔAUROC and negative ΔBrier indicated improved performance. For the transcriptomic-representation analysis, performance of both *C* + *B* + *K* and *C* + *B* + elastic-net-selected genes was evaluated relative to the common *C* + *B* baseline using the same paired-difference convention, outer CV partitions, and performance metrics. All metrics and paired performance differences were summarized across the 10 repeats using the mean and standard deviation.

### Feature Importance

Feature importance was assessed using ridge logistic regression coefficients from the *C* + *B* and *C* + *B* + *K* models fit on all 298 response-evaluable IMvigor210 patients. Coefficient sign indicated the direction of association with response, and absolute magnitude was used to describe relative model contribution. Coefficients were compared between models to assess how adding Block *K* altered the weighting of clinical and biomarker predictors. To further assess the contribution of individual Block *K* features, an ablation study was performed. Each of the eight TME gene-set scores was removed one at a time from the *C* + *B* + *K* model, and performance was reevaluated using the same CV partitions as the complete model. The ablation effect was defined as ΔAUPRC_ablation_ = AUPRC_*C* + *B* + *K*_ − AUPRC_*C* + *B* + *K* − feature_. Positive values indicate that removing the feature reduced model performance.

### Sensitivity Analysis

To address the known ambiguity of ICI response definition, patients with stable disease (SD) were excluded from non-responders, and CR/PR was compared directly with PD to assess the contribution of knowledge-guided TME features under this alternative, more distinct radiographic response phenotype definition. The repeated nested CV procedure was rerun in the reduced cohort using newly generated stratified partitions.

### External Validation

External validation was evaluated in GSE176307, an independent cohort of patients with metastatic urothelial carcinoma treated with PD-1 or PD-L1 inhibitors17. After matching clinical and RNA-sequencing records and excluding patients without an evaluable response, 87 patients were included. Because several IMvigor210 predictors were unavailable in GSE176307, evaluation used a harmonized feature set shared across cohorts. The harmonized clinical block included sex, ECOG performance status, and tobacco use history; ECOG was categorized as 0, 1, or ≥ 2, and light smokers were grouped with current smokers. The biomarker block included TMB and *CD274* mRNA expression; *CD274* expression was used in both cohorts because comparable PD-L1 IHC measurements were unavailable externally. Knowledge-guided transcriptomic scores were calculated identically using genes available in both datasets.

External comparisons included *C* vs. *C* + *K* and *C* + *B* vs. *C* + *B* + *K*. Block *R* was not evaluated because neoantigen burden and immune phenotype were unavailable in GSE176307. For each comparison, ridge regularization was selected using five-fold CV in IMvigor210. The model and preprocessing pipeline were then refit on all 298 response-evaluable IMvigor210 patients and applied without modification to GSE176307. No model fitting or hyperparameter tuning was performed using the external cohort. External performance was evaluated using AUPRC, AUROC, and Brier score. 95% confidence intervals (CIs) for each metric were estimated using 1,000 patient-level bootstrap resamples, with paired resampling used for ΔAUPRC, ΔAUROC, and ΔBrier. Sensitivity analyses excluded patients with original ECOG 3, light smoking status, or both, without retraining the models.

## Results

### Dataset and Study Cohort

Among 348 patients in IMvigor210, 298 had an evaluable response and were included in response modeling. Responses included 25 CR, 43 PR, 63 SD, and 167 PD. Overall, 68 patients were classified as responders and 230 as non-responders, corresponding to a response rate of 22.8%. Baseline clinical and biomarker characteristics of the IMvigor210 cohort are summarized in Table 1.

**Table 1.**
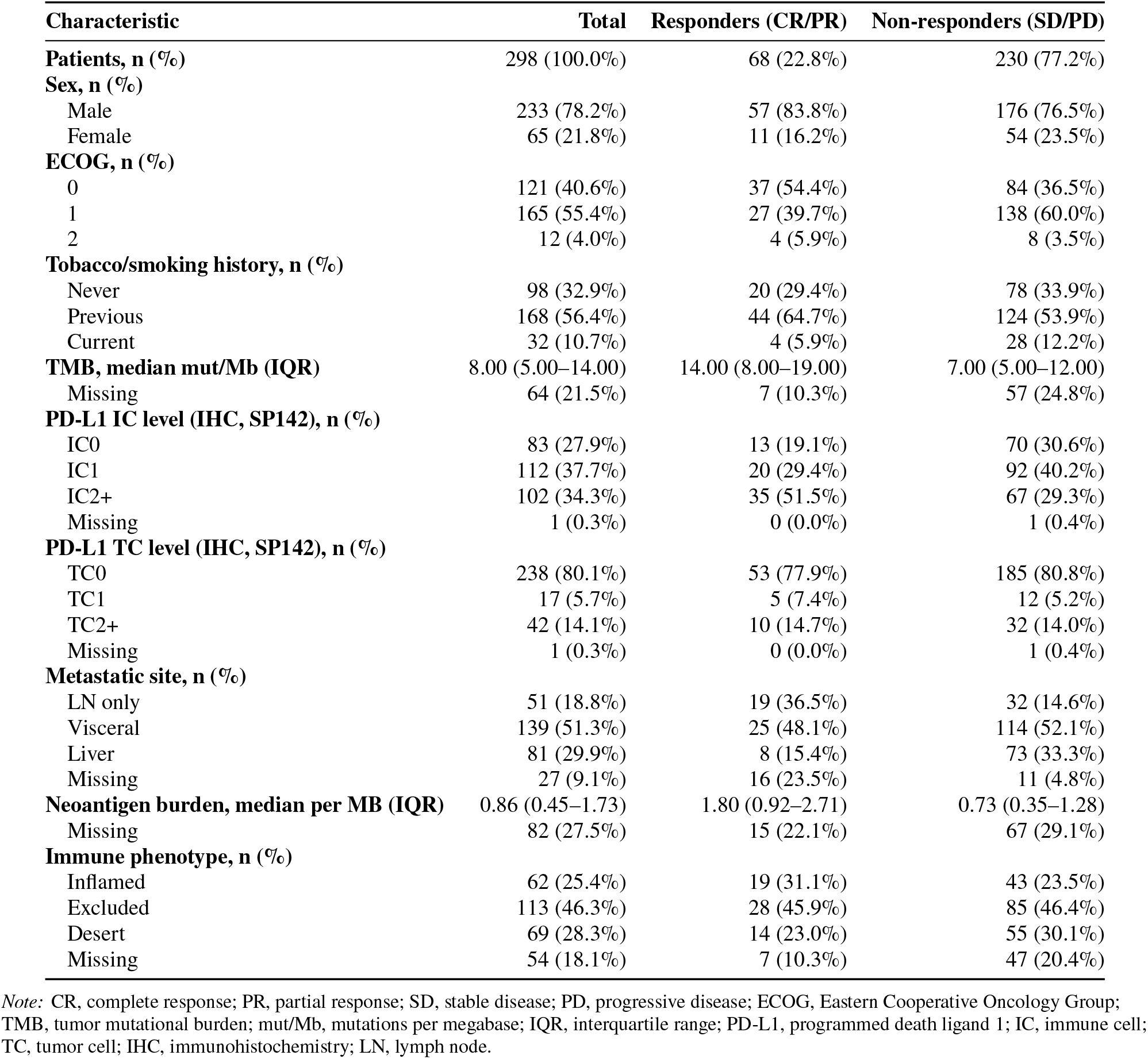
Baseline characteristics of the IMvigor210 training cohort.

### Transcriptomic Feature Engineering

#### Knowledge-Guided TME Gene Sets

CD8 T-effector and cytotoxic lymphocyte scores were higher in responders than non-responders and remained significant after BH-FDR correction (both *q* = 0.020). Median CD8 T-effector scores were 3.29 in responders and 2.76 in non-responders (Δ median = +0.53), while cytotoxic lymphocyte scores were 1.85 and 1.42, respectively (Δ median = +0.43). Endothelial-cell scores were lower in responders at the nominal level (*p* = 0.037) but did not remain significant after FDR correction (*q* = 0.099). No other gene-set comparison met the FDR threshold (Table 2).

**Table 2.** Comparison of knowledge-guided TME gene-set scores between responders and non-responders.

| Signature | Median (Responders) | Median (Non-responders) | $\Delta$ Median | $p$-value | $q$-value (FDR, $n = 8$) |
| --- | --- | --- | --- | --- | --- |
| CD8 T-effector | 3.29 | 2.76 | +0.53 | 0.005 | **0.020** |
| Cytotoxic lymphocytes | 1.85 | 1.42 | +0.43 | 0.005 | **0.020** |
| Endothelial cells | 2.61 | 2.71 | -0.11 | 0.037 | 0.099 |
| APM | 8.76 | 8.29 | +0.47 | 0.055 | 0.108 |
| NK cells | 0.37 | 0.28 | +0.09 | 0.068 | 0.108 |
| Fibroblasts | 7.09 | 7.43 | -0.34 | 0.158 | 0.183 |
| Immune checkpoint | 1.87 | 1.63 | +0.24 | 0.160 | 0.183 |
| Monocytic lineage | 3.93 | 3.87 | +0.06 | 0.956 | 0.956 |

Note: $\Delta$ median was calculated as the median signature score in responders (CR/PR) minus that in non-responders (SD/PD). Group differences were assessed using two-sided Mann-Whitney U tests. $q$ -values were calculated using the Benjamini-Hochberg false discovery rate correction across eight prespecified signatures. Bold values indicate FDR-adjusted $q < 0.05$ .

#### Data-Driven Feature Selection

After variance filtering, a mean of 18,297 genes remained eligible for selection across outer folds (range, 18,225–18,357), of which elastic net selected a mean of 363 genes per outer fold (range, 139–544). Across all 50 outer folds, 2,860 unique genes were selected at least once; 182 were selected in at least 50% of folds, 49 in at least 80%, and 26 in at least 90%. Gene selection showed modest stability, with a mean pairwise Jaccard similarity of 0.227, indicating that two fold-specific selected-gene sets shared, on average, about 23% of the genes in their combined union. Four genes were selected in all 50 outer folds: *IFNG* (interferon-γ), *KLRC2* (NKG2C, an activating NK-cell receptor), *CHST3* (involved in chondroitin sulfate biosynthesis), and *LOC102723352* (an uncharacterized locus). Among the 26 genes selected in at least 90% of folds, four overlapped the prespecified Block *K* gene sets: *IFNG* and *CXCL9* from the CD8 T-effector set, *KLRC3* from the cytotoxic lymphocyte set, and *KIR2DL4* from the NK-cell set, indicating convergence between the data-driven and knowledge-guided approaches on immune-effector biology.

Formal pathway enrichment was sparse, with IL-6/JAK/STAT3 signaling reaching BH-FDR *q* < 0.05 in only 1 of 50 outer folds. Based on pathway rank recurrence across folds, IL-6/JAK/STAT3 signaling appeared among the five pathways with the smallest enrichment *p*-values in 35 of 50 folds (70%), followed by angiogenesis (44%) and genes downregulated by KRAS activation (42%) (Table 3).

**Table 3.**
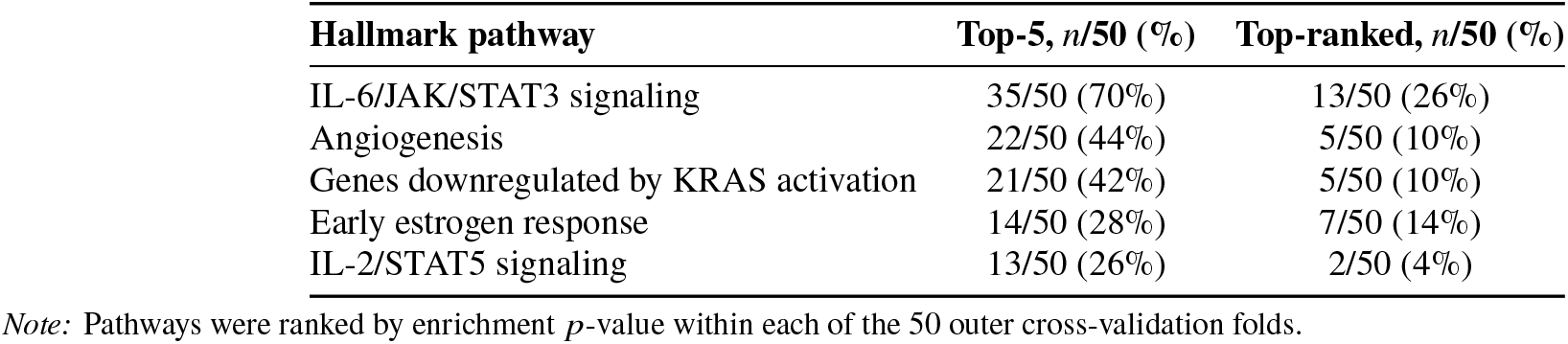
Recurrence of top-ranked Hallmark pathways.

### Model Evaluation

Across the three baselines, the largest improvement from Block *K* occurred when only clinical variables were available (Table 4). For *C* vs. *C* + *K*, AUPRC increased from 0.418 ± 0.015 to 0.476 ± 0.021 (ΔAUPRC = +0.058 ± 0.028), accompanied by higher AUROC and lower Brier score. In the prespecified primary comparison, *C* + *B* + *K* achieved an AUPRC of 0.536 ± 0.025 compared with 0.525 ± 0.029 for *C* + *B* (ΔAUPRC = +0.011 ± 0.027); AUROC was essentially unchanged, while Brier score decreased from 0.149 to 0.145. Adding Block *K* to *C* + *B* + *R* produced a similarly modest increase in AUPRC (ΔAUPRC = +0.016 ± 0.031). Thus, the incremental predictive contribution of Block *K* was substantially larger against the clinical-only baseline than when tumor biomarkers were already included.

**Table 4.** Incremental predictive value of Block *K* across the biomarker-richness gradient.

| Comparison | Metric | Baseline | +K | $\Delta$ |
| --- | --- | --- | --- | --- |
| *C* vs. *C* + *K* | AUPRC | $0.418 \pm 0.015$ | $0.476 \pm 0.021$ | $+0.058 \pm 0.028$ |
| | AUROC | $0.687 \pm 0.010$ | $0.711 \pm 0.015$ | $+0.024 \pm 0.012$ |
| | Brier | $0.161 \pm 0.003$ | $0.155 \pm 0.004$ | $-0.006 \pm 0.005$ |
| *C* + *B* vs. *C* + *B* + *K* | AUPRC | $0.525 \pm 0.029$ | $0.536 \pm 0.025$ | $+0.011 \pm 0.027$ |
| | AUROC | $0.761 \pm 0.019$ | $0.758 \pm 0.019$ | $-0.003 \pm 0.015$ |
| | Brier | $0.149 \pm 0.005$ | $0.145 \pm 0.004$ | $-0.003 \pm 0.005$ |
| *C* + *B* + *R* vs. *C* + *B* + *R* + *K* | AUPRC | $0.511 \pm 0.026$ | $0.527 \pm 0.031$ | $+0.016 \pm 0.031$ |
| | AUROC | $0.767 \pm 0.026$ | $0.755 \pm 0.026$ | $-0.012 \pm 0.031$ |
| | Brier | $0.152 \pm 0.007$ | $0.147 \pm 0.005$ | $-0.005 \pm 0.008$ |

Note: *C*, clinical variables; *B*, standard tumor biomarkers; *R*, research-enriched biomarkers; *K*, knowledge-guided TME gene-set features. Values are mean ± standard deviation across 10 repeats of outer five-fold cross-validation. $\Delta$ denotes the metric for the model including Block *K* minus the corresponding baseline model.

Under the primary *C* + *B* baseline, predictive performance differed substantially by transcriptomic representation (Table 5). *C* + *B* + *K* achieved an AUPRC of 0.536 ± 0.025, compared with 0.386 ± 0.035 for *C B* elastic-net-selected genes. The elastic-net-selected gene model underperformed the *C* + *B* baseline in all 10 repeats (mean ΔAUPRC = −0.140 ± 0.052) and also had lower AUROC (0.671 ± 0.021) and higher Brier score (0.207 ± 0.010). In contrast, the knowledge-guided representation maintained performance similar to or modestly better than the *C* + *B* baseline across the evaluated metrics.

**Table 5.** Comparison of knowledge-guided and data-driven transcriptomic representations.

| Model | AUPRC | AUROC | Brier score |
| --- | --- | --- | --- |
| $C + B$ | $0.525 \pm 0.029$ | **$0.761 \pm 0.019$** | $0.149 \pm 0.005$ |
| $C + B + K$ | **$0.536 \pm 0.025$** | $0.758 \pm 0.019$ | **$0.145 \pm 0.004$** |
| $C + B$+ elastic-net genes | $0.386 \pm 0.035$ | $0.671 \pm 0.021$ | $0.207 \pm 0.010$ |

*Note:* $C$ , clinical variables; $B$ , standard tumor biomarkers; $K$ , knowledge-guided TME gene-set features. Elastic-net genes denote transcriptome-wide genes selected within each outer training fold. Values are mean ± standard deviation across 10 repeats of outer five-fold cross-validation. Bold indicates the most favorable point estimate for each metric.

### Feature Importance

TMB had the largest positive coefficient in both the *C* + *B* and *C* + *B* + *K* models (Figure 1). The five features with the largest coefficient magnitudes were unchanged after adding Block *K*: TMB, two metastatic-site indicators, and two ECOG indicators remained the dominant predictors, suggesting that the transcriptomic features supplemented rather than displaced existing predictors. Among Block *K* features, CD8 T-effector had a prominent positive coefficient, whereas immune checkpoint and monocytic lineage had negative coefficients among the largest magnitudes.

**Figure 1.**
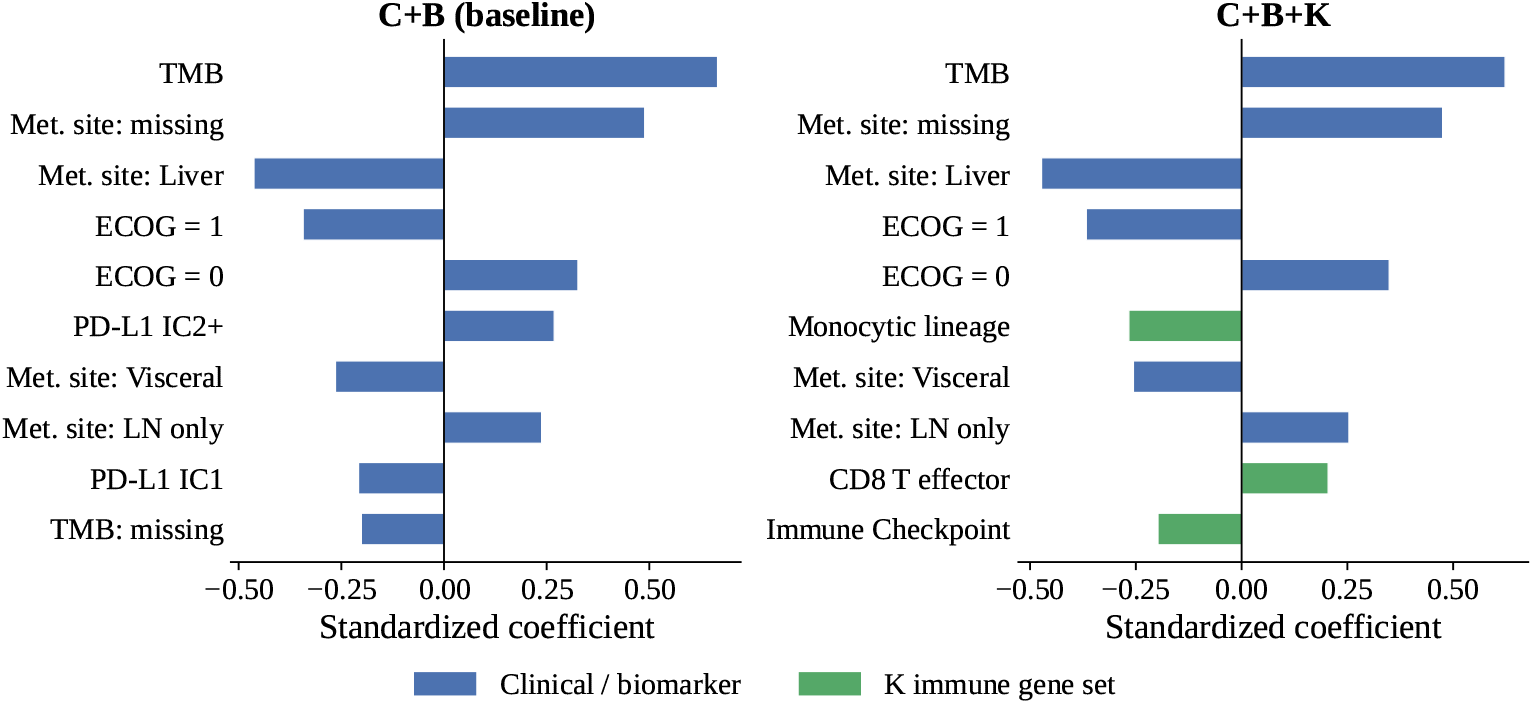
Ridge logistic regression coefficients for the top 10 features by magnitude, comparing the *C* + *B* (left) and *C* + *B* + *K* (right) models. Blue bars indicate clinical (*C*) or standard biomarker (*B*) features; green bars indicate Block *K* TME gene-set scores, present only in the *C* + *B* + *K* model. Positive coefficients indicate association with response (CR/PR); negative coefficients indicate association with non-response (SD/PD).

Consistent with the coefficient analysis, removal of CD8 T-effector produced the largest mean reduction in AUPRC in the ablation analysis (ΔAUPRC = + 0.007) and reduced performance in 7 of 10 repeats. Ablation effects for the remaining gene-set scores were small and inconsistent, indicating that no single Block *K* feature dominated its predictive contribution.

### Sensitivity Analysis

After excluding patients with SD, the incremental value of Block *K* remained positive across all three feature baselines. Mean ΔAUPRC was 0.087 for *C* vs. *C* + *K*, +0.041 for *C* + *B* vs. *C* + *B* + *K*, and 0.020 for *C* + *B* + *R* vs. *C* + *B* + *R* + *K*. Gains remained greatest when only clinical variables were available and decreased as additional biomarker information was incorporated. Compared with the primary CR/PR vs. SD/PD analysis, the incremental contribution of Block *K* was consistently larger after restricting the outcome to CR/PR vs. PD, particularly for the *C* and *C* + *B* baselines (Figure 2).

**Figure 2.**
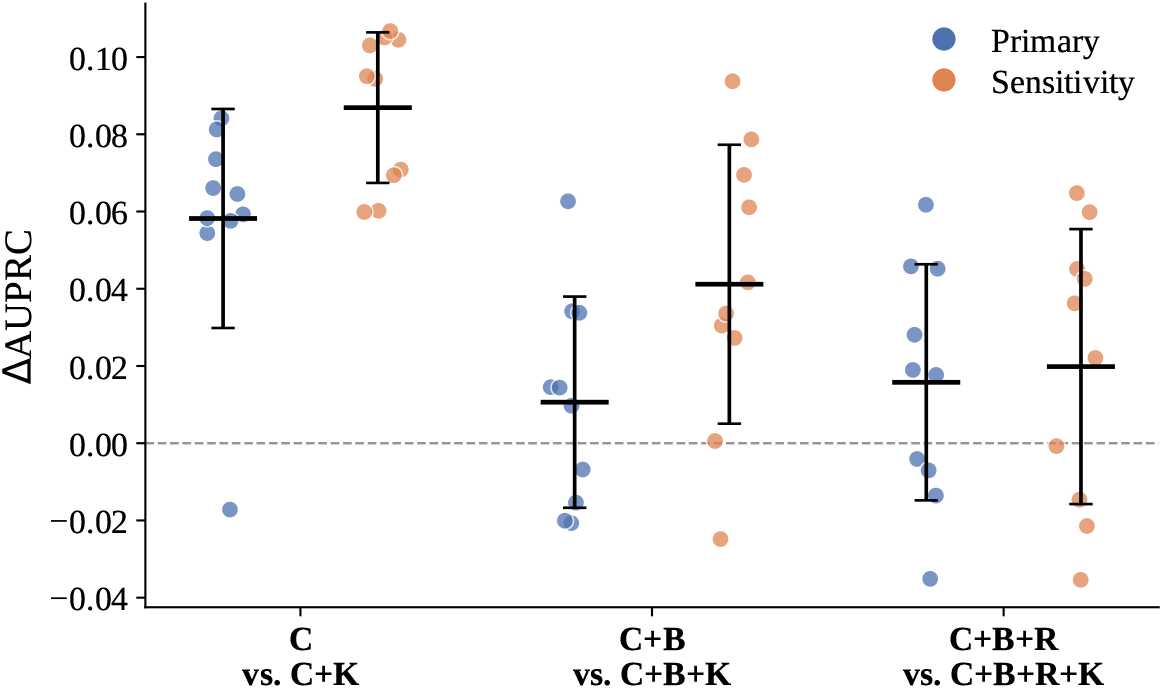
ΔAUPRC across the biomarker-richness gradient, primary (CR/PR vs. SD/PD) and sensitivity (CR/PR vs. PD) analyses. Each point shows one of 10 repeats of outer five-fold CV; bars show mean ± SD across the 10 repeats.

### External Validation

The external cohort included 87 response-evaluable patients with matched clinical and transcriptomic data. Of these, 16 patients (18.4%) were responders (7 CR and 9 PR) and 71 were non-responders (4 SD and 67 PD). Compared with IMvigor210, the external cohort included a higher proportion of female patients (37.9% vs. 21.8%) and had a lower median tumor mutational burden (5.3 vs. 8.0 mutations/Mb). Patients also received multiple PD-1/PD-L1 inhibitors, including pembrolizumab (52.9%), atezolizumab (37.9%), nivolumab (5.7%), durvalumab (2.3%), and avelumab (1.1%), rather than atezolizumab alone, providing a clinically distinct cohort for evaluating model transportability.

External evaluation reproduced the larger incremental contribution of Block *K* at the clinical-only baseline (Table 6). For *C* vs. *C* + *K*, AUPRC increased from 0.174 to 0.301 (ΔAUPRC = +0.128, 95% CI 0.011 to 0.294), while AUROC increased from 0.450 to 0.613 (ΔAUROC = +0.162, 95% CI 0.031 to 0.291). Brier score decreased from 0.155 to 0.148, although the confidence interval for the paired difference included zero. In the *C* + *B* comparison, adding Block *K* produced only small changes in performance: AUPRC increased from 0.414 to 0.434 (ΔAUPRC = + 0.019, 95% CI − 0.091 to 0.131), with minimal changes in AUROC and Brier score. Overall, the external cohort supported the internal finding that the incremental value of Block *K* was greatest when fewer non-transcriptomic biomarkers were available.

**Table 6.** External validation of the incremental predictive value of Block K.

| Comparison | Metric | Baseline (95% CI) | +*K* (95% CI) | $\Delta$ (95% CI) |
| --- | --- | --- | --- | --- |
| *C* vs. *C* + *K* | AUPRC | 0.174 (0.105–0.283) | 0.301 (0.148–0.513) | **+0.128 (0.011–0.294)** |
|  | AUROC | 0.450 (0.326–0.584) | 0.613 (0.454–0.761) | **+0.162 (0.031–0.291)** |
|  | Brier | 0.155 (0.112–0.206) | 0.148 (0.097–0.209) | -0.006 (-0.020–0.009) |
| *C* + *B* vs. *C* + *B* + *K* | AUPRC | 0.414 (0.226–0.646) | 0.434 (0.231–0.648) | +0.019 (-0.091–0.131) |
|  | AUROC | 0.743 (0.625–0.849) | 0.750 (0.623–0.858) | +0.007 (-0.104–0.118) |
|  | Brier | 0.132 (0.089–0.178) | 0.134 (0.084–0.193) | +0.003 (-0.011–0.019) |

*Note:* *C*, clinical variables; *B*, standard tumor biomarkers; *K*, knowledge-guided TME gene-set features. Values are point estimates with 95% confidence intervals from 1,000 patient-level bootstrap resamples. $\Delta$ denotes the metric for the model including Block *K* minus the corresponding baseline model. Bold $\Delta$ values indicate confidence intervals excluding zero.

Sensitivity analyses addressing ECOG and smoking-status harmonization produced the same overall pattern. After excluding patients with original ECOG 3, light smoking status, or both, ΔAUPRC for *C* vs. *C* + *K* ranged from +0.132 to +0.147, with all 95% CIs excluding zero. In contrast, ΔAUPRC for *C* + *B* vs. *C* + *B* + *K* remained small, ranging from +0.016 to +0.032, with all 95% CIs including zero.

## Discussion

Using the IMvigor210 cohort of patients with advanced urothelial carcinoma treated with atezolizumab, we evaluated whether pretreatment transcriptomic features improve prediction of response beyond clinical characteristics and tumor biomarkers, and whether transcriptomic representation influences predictive performance. Knowledge-guided TME features captured meaningful response-related biology, but their incremental predictive value was modest once tumor biomarkers were available. Their contribution was greater when prediction relied primarily on clinical variables, and the knowledge-guided TME representation was more stable and predictive than data-driven transcriptome-wide gene selection. The same general pattern was observed in an independent immunotherapy-treated cohort. Together, these findings show that the value of transcriptomic profiling depends on both the information already available and the computational representation to incorporate the molecular data.

The biological patterns observed in our analysis were consistent with established correlates of ICI response. CD8 T-effector and cytotoxic lymphocyte scores were higher in responders, while TMB remained a prominent predictor after transcriptomic features were incorporated, aligning with prior IMvigor210 findings linking T-cell effector activity and tumor mutational features to atezolizumab response^4^. However, biological relevance did not translate into substantial incremental predictive value once tumor biomarkers were available. TMB and PD-L1 already capture aspects of tumor immunogenicity and an inflamed TME, while Block *R* further incorporated neoantigen burden and immune phenotype, resulting in considerable biological overlap with the processes captured by the knowledge-guided transcriptomic scores^6^. Missingness in these research-enriched biomarkers may also have limited their contribution. Thus, the modest gain from transcriptomics in biomarker-rich models likely reflects overlapping biological information and incomplete measurement of related tumor-immune features.

The contrast between knowledge-guided and data-driven transcriptomic representations highlights a common challenge in high-dimensional clinical genomics: extracting reliable predictive information when molecular measurements greatly outnumber patients, a setting in which gene-level signatures can be highly sensitive to sampling variation^9^. Elastic net repeatedly selected biologically plausible immune genes, including *IFNG* and *CXCL9*, yet the selected genes varied substantially across training samples and yielded poorer prediction than the knowledge-guided representation. Knowledge-guided aggregation may reduce this instability by summarizing coordinated biological processes rather than relying on individual transcripts. Despite gene-level variability, pathway analysis suggested convergence at a broader biological level: IL-6/JAK/STAT3 signaling was the most recurrent Hallmark pathway. IL-6/STAT3 signaling has been linked to resistance to PD-L1 blockade and impaired CD8 T-cell cytotoxic differentiation, providing a plausible biological connection to the immune-effector signals observed in our analysis^18^.

The larger incremental contribution of Block *K* after excluding SD may reflect the clinical heterogeneity of stable disease during immune checkpoint blockade. SD can represent durable disease control, indolent tumor behavior, or early treatment resistance, and prior studies have shown substantial outcome heterogeneity within this group^19, 20^. Restricting the analysis to CR/PR vs. PD therefore created more distinct response phenotypes, which may have made transcriptomic differences easier to detect. This finding also highlights the importance of outcome definition in immunotherapy prediction studies, where heterogeneous intermediate response states can attenuate predictive signal.

External validation provided additional support for the main findings. Although GSE176307 differed from IMvigor210 in patient characteristics and included several PD-1/PD-L1 inhibitors rather than atezolizumab alone^17^, the same relative pattern was observed: knowledge-guided transcriptomic features contributed more when added to clinical variables alone and little once the harmonized tumor biomarker block was included. Sensitivity analyses excluding patients affected by the ECOG and smoking-status harmonization decisions produced similar results, suggesting that these specific mapping choices did not drive the observed pattern. This consistency supports the transportability of the overall predictive pattern, even though absolute model performance differed across cohorts.

Several limitations should be considered. First, the modest sample size, particularly the limited number of responders, constrained high-dimensional feature discovery and likely contributed to the unstable gene-level selection. Second, bulk RNA sequencing averages expression across heterogeneous tumor and microenvironmental cell populations and does not preserve spatial organization, which may obscure response-relevant cellular states and interactions. Third, the external cohort was small, required feature harmonization across cohorts, and supported validation of only a subset of the internal modeling framework, thus limiting the precision and scope of transportability assessment.

More broadly, these findings suggest that molecular profiling should be judged by the incremental value it provides beyond information already available in clinical practice. As sequencing becomes increasingly accessible, transcriptomic profiling could refine treatment selection when established biomarkers are insufficient, with the broader goal of advancing precision immuno-oncology. In smaller clinical cohorts, incorporating prior biological knowledge may provide a more reliable way to use these high-dimensional data. Future studies should evaluate this approach prospectively and determine whether spatial, single-cell, or other molecular measurements provide additional value.

## Conclusion

Knowledge-guided TME transcriptomic features added predictive information for immunotherapy response, but their incremental value depended on the information already available: gains were greatest with clinical variables alone and diminished as tumor biomarkers were included. In a modest-sized cohort, biologically guided TME gene-set representation was also more stable and predictive than data-driven transcriptome-wide gene selection, with the same relative pattern observed in an independent cohort. As sequencing becomes increasingly available in clinical oncology, its value will depend on whether molecular data add meaningful predictive information beyond existing biomarkers, and whether that information can be translated into reliable tools for more precise treatment selection.

## Data Availability

All data produced in the present work are contained in the manuscript.

